# Digital divide is amplifying cognitive disparities among the older population: a community-based cohort study in China

**DOI:** 10.1101/2024.02.24.24303323

**Authors:** Yumeng Li, Chen Liu, Jiaqing Sun, Junying Zhang, Xin Li, Zhanjun Zhang

**Author notes:** Correspondence to: Xin Li, Ph.D., State Key Laboratory of Cognitive Neuroscience and Learning, Beijing Normal University, Beijing 100875, China; Email addresses, Zhanjun Zhang, Ph.D., State Key Laboratory of Cognitive Neuroscience and Learning, Beijing Normal University, Beijing 100875, China; Email addresses. **Statement of Authors’ Contributions** LYM and LX conceived the study. LYM and LC participated in the data collection. LYM carried out the statistical analysis. LYM and LX analyzed and interpreted the data. LYM wrote the manuscript. LX and SJQ revised the manuscript. ZZJ and ZJY supervised and coordinated the study. All authors contributed to the improvement of this manuscript and approved the final version for submission.

## Abstract

**Objectives:** To investigate the potential impact of the digital divide on individuals’ cognitive function and its association with the development and reversion of mild cognitive impairment (MCI).

**Methods:** This cohort study used data from Beijing Aging Brain Rejuvenation (BABRI) study applying a multistage cluster sampling design in 2008-2020. Analysis of Covariance (ANCOVA), mixed linear models, and Cox proportional hazards models were used to model the association of digital divide and multi-domain cognition.

**Results:** Among the 10098 participants, nearly half of them (48.9%) failed to overcome the digital divide, resulting in the worse performance in processing speed (F=10.67, *p*<0.001). The causal model indicated that individuals’ physical and mental health joint educational and occupational prestige affected the resource they achieved ultimately caused the digital divide. Moreover, longitudinal data revealed that both the elderly who successfully crossed the digital divide during the tracking process and those who had already done so prior to tracking showed significantly slower rates of decline in processing speed (B=-1.98, *p*<0.05; B=-2.62, *p*<0.01) and general cognitive function (B=3.50, *p*<0.001; B=3.13, *p*<0.01). Additionally, overcoming the digital divide also exhibited a lower risk of developing into MCI (HR, 0.5; 95% CI, 0.34-0.74; HR, 0.43; 95% CI, 0.29-0.62) and a greater probability of reversion from MCI to normal cognition (HR, 6.00; 95% CI, 3.77-9.56; HR, 9.22; 95% CI, 5.63-15.11).

**Conclusions:** Overcoming the digital divide was significantly associated with better performance and lower aging rate of cognitive function, as well as a lower risk of developing into MCI and a higher probability of reversion from MCI into NC.

## Introduction

In today’s modern society, digital information and communication technologies (ICTs) have become indispensable in our daily lives. The ‘digital divide’ has become a vivid metaphor to describe the perceived inequality of those who either are unable to make use of ICTs. ^1,2^

The Digital Divide (DD) exerts a profound impact on the older population, which is experiencing rapid growth in numerous countries, including China, where both absolute numbers and relative proportions are continuously increasing. This demographic exhibit significantly lower rates of acceptance and utilization of ICTs, leading to an increasing trend of exclusion among the elderly in the dynamically advancing information society.^3–5^

The understanding of this phenomenon benefits from the causal model of resources and appropriation theory of the digital divide.^2,6,7^ The core hypothesis of this theory posits that disparities in personal characteristics (such as health and habits) and positional characteristics (such as education and occupation) contribute to unequal access to diverse resources (such as mental well-being and social relationships), thereby resulting in the digital divide. However, there is currently a scarcity of empirical studies that comprehensively incorporate various influential factors and investigate the causal pathways underlying the digital divide.

The digital divide can potentially contribute to the adoption of a less engaging lifestyle throughout the lifespan of the elderly, which may diminish their “cognitive reserve” (the resource of the brain can get to tolerate the age-related changes or pathology related to dementia^8^) and ultimately hasten the deterioration of cognitive function.^9^ The evidence suggested that daily internet use was associated a reduced risk of mild cognitive impairment (MCI).^10^ Also, longitudinal research indicated that overcoming the digital divide was associated with a decline in the incidence of MCI among older adults after a period of ten years.^11^ Nonetheless, existing literature focuses too narrowly on cognitive domains and fails to consider the specificity of cognitive differences caused by digital divide. Similarly, the longitudinal changes in multi-domain cognitive function influenced by the digital divide have not been adequately considered. Additionally, while many studies have investigated the progression of MCI, some have found that a considerable proportion, ranging from 24% to 50%, of individuals with MCI may actually revert back to a normal cognition (NC) status.^12–14^ Thus, it is crucial to take into account the digital divide influences both the development and reversion of MCI.

It can be foreseen that the digital divide will put the older population in a state of resource inequality, which will have an impact on their future cognitive development. Based on the large-scale database, the current research will investigate the following aspects: (1) the potential causal mechanism of multifarious influential factors contributing to unequal resource distribution in relation to the digital divide; (2) the cross-sectional differences and longitudinal aging rate alternations of multi-domain cognitive function caused by digital divide; (3) the associations of digital divide with the development and reversion of MCI while excluding the normal risk factors.

## Methods

### 1 Study Participants

The BABRI cohort study is based on the registry of a large community population in Beijing, which collects comprehensive information on aging, tracks changes in cognitive function over years.^15^ All of the participants were 50 years or above at the time of base line enrollment, capable of living independently, without nervous system diseases or psychiatric disorders, having 6 or more years of formal education, which is required for the cognitive assessments. By applying a multistage cluster sampling design, the recruitment of participants was mainly conducted in the communities in Beijing, from 2008 to 2019. A total of 10098 qualified individuals were recruited from the communities of these districts. All of the participants who were registered in BABRI would be revisited every 2 or 3 years within the total duration of the 20 years of the project. This study will exclude longitudinal participants in the next phase of analysis based on the following criteria: (1) clinical diagnoses of neurodegenerative diseases (e.g., AD and Parkinson’s disease), serious brain diseases (e.g., severe cerebrovascular diseases, brain tumors, and brain trauma), or psychiatric disorders (e.g., severe major depression disease, bipolar disorder, and schizophrenia); (2) histories of substance or alcohol abuse/dependence; (3) The assessment indicators for the digital divide are missing in the tracking process.

### 2 Measurements

The Resources and Appropriation Theory of the digital divide was developed over a ten-year period, culminating in its full and mature presentation.^7^ The core hypothesis of this theory illustrated that individual’s characteristics and status leads to unequal access to resources they are able to achieve, ultimately resulting in the digital divide. In this study, the influencing factors of digital divide were classified into three main parts. The independent variables are personal categories and positional categories, while the mediation variable is resources.

#### 2.1 Personal categories

Personal categories encompass various factors, such as physical health (subjective health status, BMI and chronic disease), mental health (the geriatric depression scale and FACE), smoking, and alcohol consumption.

##### 2.1.1 Physical health: the subjective health status (SHS), BMI index and chronic diseases

SHS refers to a question: “Please provide an overall evaluation of your health status: is it good, fair, poor, or very poor”. Body mass index(BMI) was calculated by dividing their weight in kilograms by their height in meters squared. Medical history of individuals was inquired regarding hypertension, diabetes and hyperlipidemia and this information was also validated with the diagnosis and management records from community health service institutions.

##### 2.1.2 Mental health

The geriatric depression scale (GDS) is a standardized depression scale for older adults.^16,17^ The FACE scale was used to measure the subjective well-being(SWB) The higher scores the older adults get, the less SWB they feel.^18^

##### 2.1.3 Daily unhealthy habits

The smoke and alcohol consumption are binary variables indicating whether the individual smoke or consume alcohol every day.

#### 2.2 Positional categories

Positional categories include the variables manifesting the individuals’ social status, such as educational status, subjective social status and occupational prestige. The educational status refers to individual’s the specific duration of education. The subjective social status refers to a question: “How would you evaluate your current socio-economic status?” The response is a four-point rating namely rich, upper-middle, lower-middle, and poor. The occupational prestige is measured by the occupational prestige scale based on the Chinese national survey data.^19^

#### 2.3 Resources

The resources include the economic resources (income range individual could achieve), mental resources and social resources. The economic resources refer to individual’s current monthly income, with 500 as the unit, divided into 13 levels. The mental and social resources are obtained by weighted summation based on the relevant items derived from the Leisure Activity Scale (LAS) which is a five-point scale that consists of 23 sub-items.^20,21^

#### 2.4 Quantifying Digital Divide

The quantification indicator of digital divide involves an item about the frequency of using ICT from LAS: “How often do you use computer and mobile”.^22–28^ We classify individuals with scores of 0(Never), 1(≥Once a year), and 2(≥Once a month) into the DD group, and individuals with scores of 4(≥Once a week) and 5(Everyday) into the ODD group. The DD group means failing to overcome the digital divide while the ODD group means overcoming the digital divide.

For cross-sectional data, participants were only classified into DD group and ODD group. In order to investigate the impact of ODD on cognitive aging, for longitudinal data participants were categorized into three groups. Two of these groups represented participants who remained in the DD or ODD group, while the third group was the Trans group. This group included participants who transitioned from the DD group to the ODD group during the tracking process. We excluded participants who transitioned from the ODD to the DD group because crossing the digital divide is a relatively stable state. If a transition occurs from ODD to DD, it may be due to uncontrollable external factors, which are not the focus of this study.

#### 2.5 Cognitive

All participants underwent a battery of neuropsychological tests at the baseline recruitment.^15^ The assessment involved in the general cognitive ability and cognitive function across five domains including memory, language, attention, visuospatial abilities, and executive function. The general cognitive ability was tested using the Chinese version of Mini-Mental State Examination (MMSE).^29^ Memory was tested using the Auditory Verbal Learning Test (AVLT)^30^ and the Rey-Osterrich Complex Figure Test (ROCF).^31^ Executive function was tested using the Stroop Color Word Test (SCWT) ^32^ and the Trail Making Test (TMT).^33^ Spatial processing was assessed using the Clock Drawing Test (CDT)^34^ and the ROCF_Copy test.^31^ Attention was evaluated using the Symbol Digit Modification Test (SDMT) ^35^ and the TMT_A test.^33^ Language was tested using the Boston Naming Test (BNT) and the Verbal Fluency Test (VFT).^36^

#### 2.6 MCI diagnostic criteria

The diagnostic criteria for Mild Cognitive Impairment (MCI) includes^37,38^: individuals without dementia, exhibiting subjective cognitive decline, and presenting with at least one cognitive domain displaying two test scores that fall below 1.5 standard deviations from the mean score of the same age and education level group in objective tests. Moreover, their general cognitive abilities and daily life functioning are typically unaffected.

#### 2.7 Demographic variables: age, gender marital status and residential status

The marital status involves three options: married, divorced, and widowed. Residential status involves three options: living alone, living with a spouse, and living with children.

### 3 Statistical Analysis

Baseline characteristics of the participants are presented as mean (SD) for continuous variables whereas categorical variables are represented as frequencies and proportions.

Z is the z value that would be obtained from performing a Manne-Whitney test The intergroup variations in terms of influential factors between the DD and ODD groups were assessed through independent samples t-test, chi-square test, and nonparametric tests. And we use Cohen‘s d and Cramer’s V for the calculation of effect size.^39–41^

An intermediary model is constructed to elucidate the impact mechanism of the digital divide, respectively using personal and positional characteristics as independent variables, resource acquisition as a mediating variable, and the variable of digital divide as the dependent variable.

The intergroup differences in cognitive abilities were examined by conducting analysis of covariance (ANCOVA) while controlling for age, education level, gender and chronic diseases. Binary logistic regression was employed to investigate the predictive impact of the DD variable on the occurrence of MCI.

A linear Bayesian change-point regression was performed on the age-group-averaged data to compare the decline trajectories of multi-domain cognition among the older population.

In order to effectively compare the impact of cognitive abilities among the DD, trans, and ODD group, dummy variable coding was applied to the longitudinal data to incorporate the effects of trans-DD and ODD-DD in the model.

The mixed linear model (MLM) was employed to examine the influence of the DD variable on the rate of cognitive aging at an individual level. Initially, we established the null model and unconditional growth model. The null model was utilized to determine the hierarchical structure of the longitudinal data for different cognitive functions, which was suitable for MLM analysis. The unconditional growth model was used to identify significant aging patterns in various cognitive functions over time. After selecting these two models, we constructed the full model that encompassed: (1) Level 1: described the individual cognitive level aging patterns, (2) Level 2: investigated the influence of the DD variable on the aging patterns of multiple cognitive abilities in individuals.

Level1:

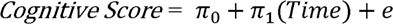

Level2:

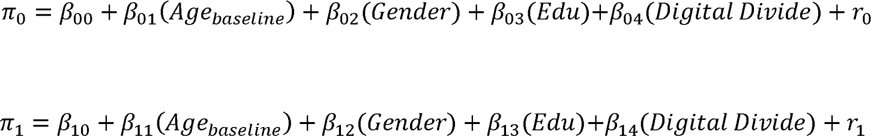

To investigate the impact of the DD variable on individual MCI development and outcomes, we employed the Cox proportional hazards model. Two models were developed, the first model focused on the progression from normal cognition to MCI, while the second model focused on the transition from MCI to normal cognition.

## 4 Results

This study included 10098 participants for the cross-sectional analysis (Age: mean[SD], 66.7[7.9] years, 6095 female[60.4%] and 4003 male[39.6%], Education: mean[SD], 10.7[3.5] years), with 4941(48.9%) belonging to DD group and 5157(51.1%) belonging to ODD group. In the longitudinal data, 2092 participants were involved. They were divided into DD group, trans group and ODD group.

Also, among these participants, their outcomes during the tracking process can be categorized into several possibilities, from normal cognition to normal cognition (NC →NC: N=1473), from the healthy to the MCI (NC→MCI, N=190), from the MCI to the MCI (MCI→MCI: N=193), from the MCI to the healthy (MCI→NC, N=201).

### 4.1 Characteristic Differences among older Population under the Digital Divide

As shown in Table 1, the personal characteristics of ODD group include having hyperlipidemia, low depression scores, high subjective well-being, and non-smoking, while the positional status of them include higher education level and higher self-perceived SES. Also, the ODD group achieve more economic, mental and social resources.

**Table 1.**
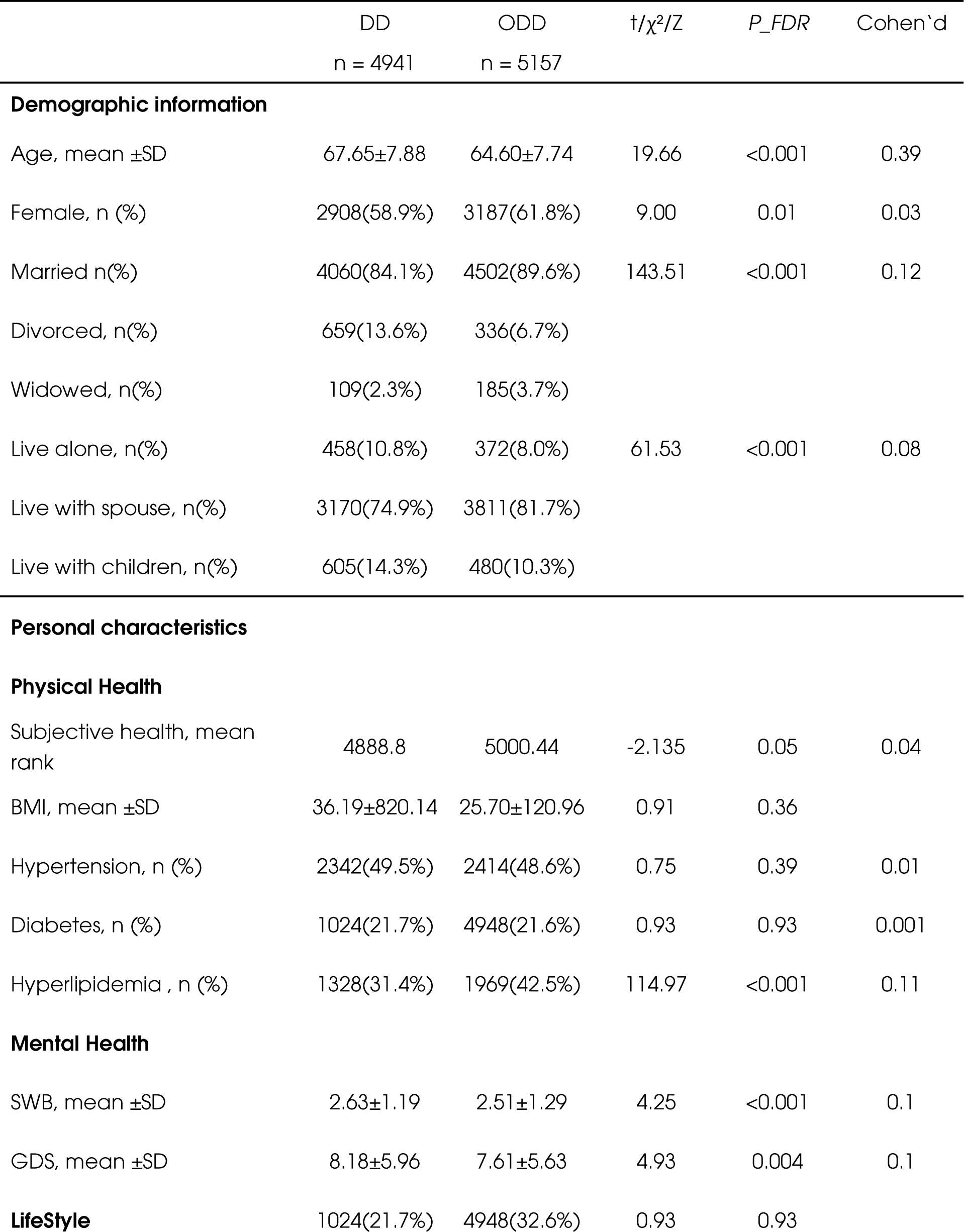
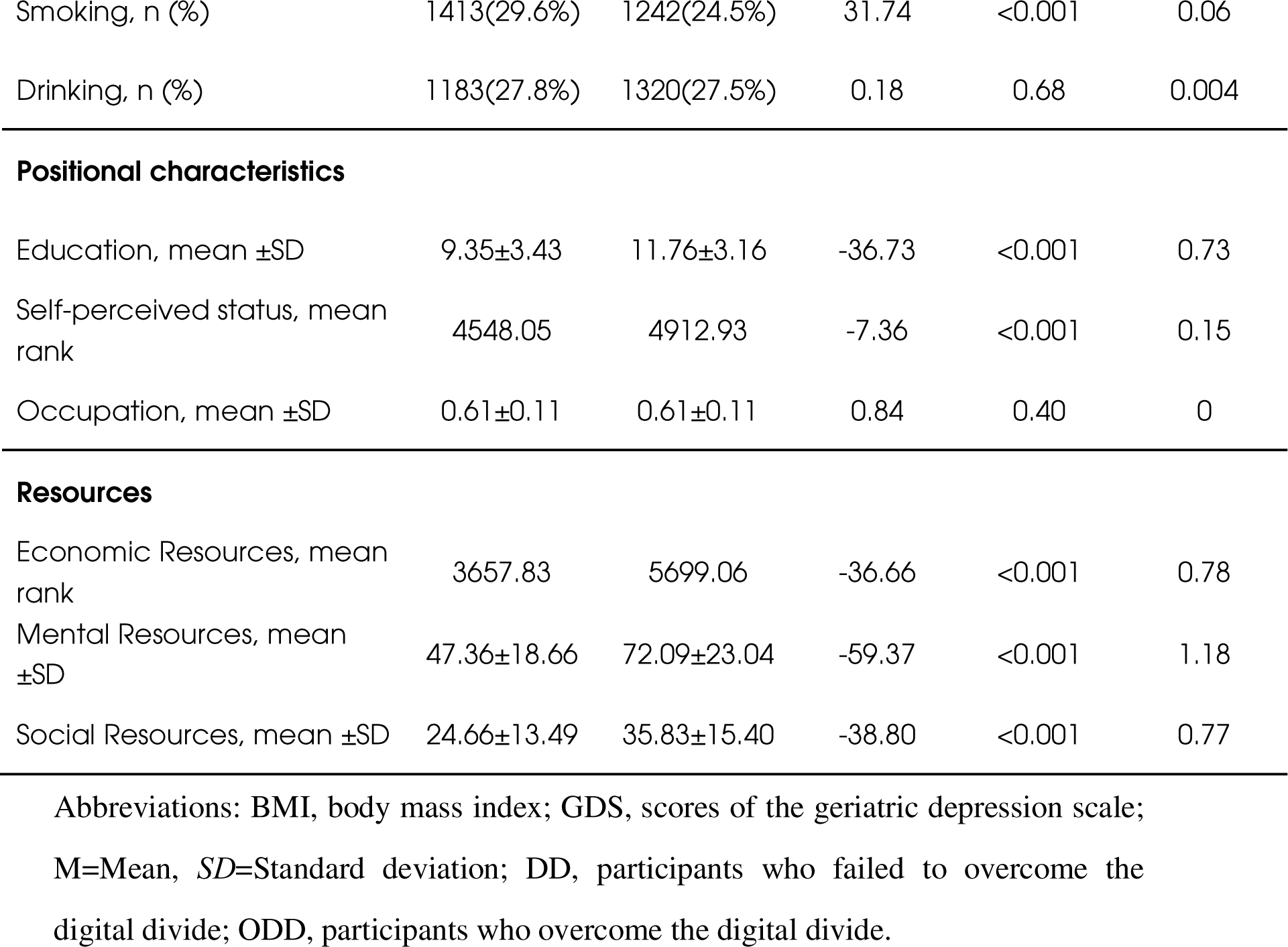
Characteristics and Digital Divide among older population.

| | DD<br>n = 4941 | ODD<br>n = 5157 | t/$\chi^2$/Z | P_FDR | Cohen'd |
| --- | --- | --- | --- | --- | --- |
| **Demographic information** |  |  |  |  |  |
| Age, mean ±SD | 67.65±7.88 | 64.60±7.74 | 19.66 | <0.001 | 0.39 |
| Female, n (%) | 2908(58.9%) | 3187(61.8%) | 9.00 | 0.01 | 0.03 |
| Married n(%) | 4060(84.1%) | 4502(89.6%) | 143.51 | <0.001 | 0.12 |
| Divorced, n(%) | 659(13.6%) | 336(6.7%) |  |  |  |
| Widowed, n(%) | 109(2.3%) | 185(3.7%) |  |  |  |
| Live alone, n(%) | 458(10.8%) | 372(8.0%) | 61.53 | <0.001 | 0.08 |
| Live with spouse, n(%) | 3170(74.9%) | 3811(81.7%) |  |  |  |
| Live with children, n(%) | 605(14.3%) | 480(10.3%) |  |  |  |
| **Personal characteristics** |  |  |  |  |  |
| **Physical Health** |  |  |  |  |  |
| Subjective health, mean rank | 4888.8 | 5000.44 | -2.135 | 0.05 | 0.04 |
| BMI, mean ±SD | 36.19±820.14 | 25.70±120.96 | 0.91 | 0.36 | |
| Hypertension, n (%) | 2342(49.5%) | 2414(48.6%) | 0.75 | 0.39 | 0.01 |
| Diabetes, n (%) | 1024(21.7%) | 4948(21.6%) | 0.93 | 0.93 | 0.001 |
| Hyperlipidemia , n (%) | 1328(31.4%) | 1969(42.5%) | 114.97 | <0.001 | 0.11 |
| **Mental Health** |  |  |  |  |  |
| SWB, mean ±SD | 2.63±1.19 | 2.51±1.29 | 4.25 | <0.001 | 0.1 |
| GDS, mean ±SD | 8.18±5.96 | 7.61±5.63 | 4.93 | 0.004 | 0.1 |
| **LifeStyle** | 1024(21.7%) | 4948(32.6%) | 0.93 | 0.93 |  |
| Smoking, n (%) | 1413(29.6%) | 1242(24.5%) | 31.74 | <0.001 | 0.06 |
| Drinking, n (%) | 1183(27.8%) | 1320(27.5%) | 0.18 | 0.68 | 0.004 |
| Positional characteristics |  |  |  |  |  |
| Education, mean ±SD | 9.35±3.43 | 11.76±3.16 | -36.73 | <0.001 | 0.73 |
| Self-perceived status, mean rank | 4548.05 | 4912.93 | -7.36 | <0.001 | 0.15 |
| Occupation, mean ±SD | 0.61±0.11 | 0.61±0.11 | 0.84 | 0.40 | 0 |
| Resources |  |  |  |  |  |
| Economic Resources, mean rank | 3657.83 | 5699.06 | -36.66 | <0.001 | 0.78 |
| Mental Resources, mean ±SD | 47.36±18.66 | 72.09±23.04 | -59.37 | <0.001 | 1.18 |
| Social Resources, mean ±SD | 24.66±13.49 | 35.83±15.40 | -38.80 | <0.001 | 0.77 |

Abbreviations: BMI, body mass index; GDS, scores of the geriatric depression scale;
M=Mean, SD=Standard deviation; DD, participants who failed to overcome the digital divide; ODD, participants who overcome the digital divide.

### 4.2 Resources mediated the personal and positional influence of Digital Divide

The mediation model in Figure 1 illustrates the potential mechanism of the digital divide formation among the elderly population, primarily due to inequalities in individual characteristics and status features that result in unequal access to economical, mental and social resources, ultimately leading to the digital divide.

**Figure 1.**
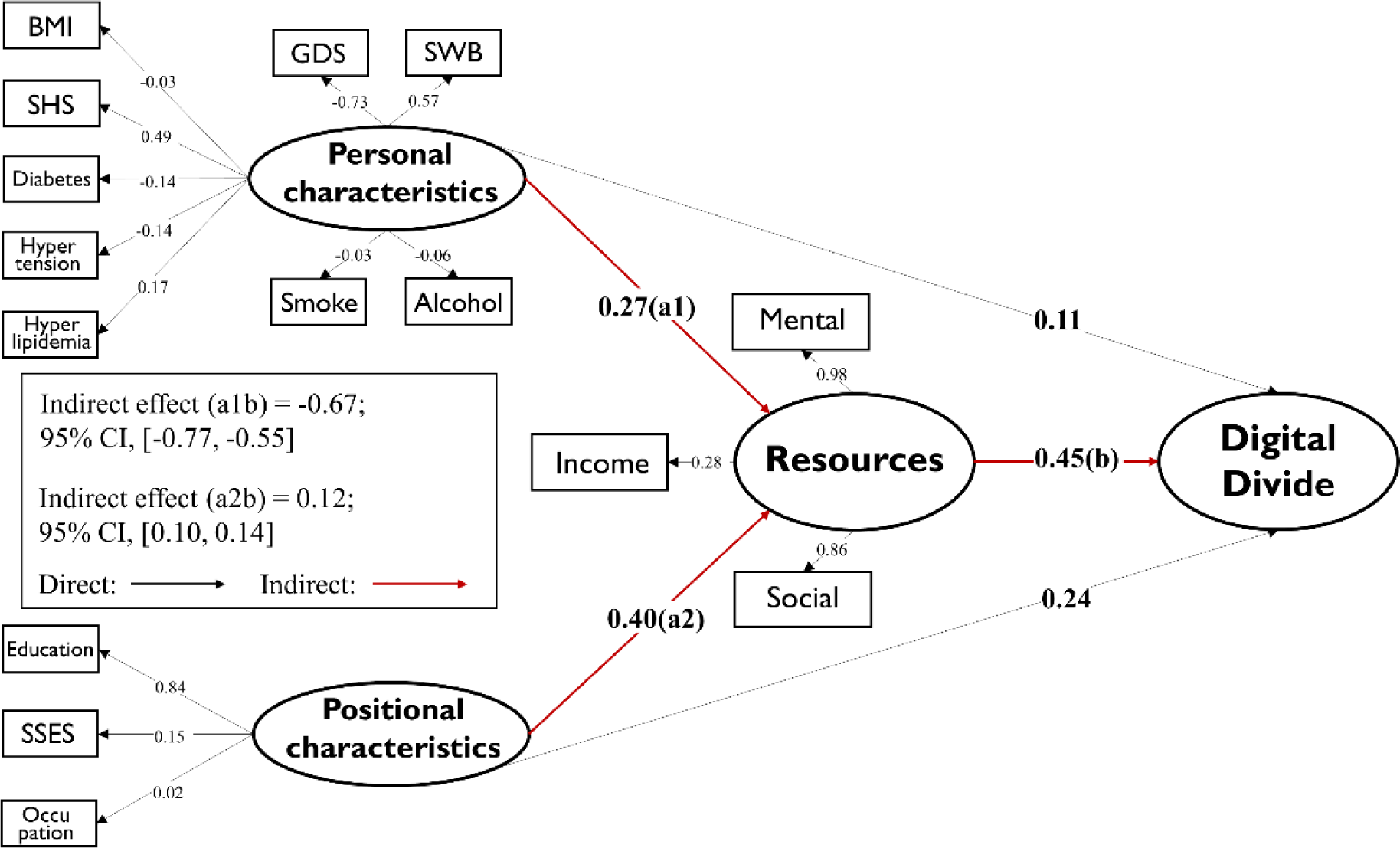
Mediation model Elucidating the Mechanism of the Digital Divide. The model is adjusted for the demographic variables (age, gender, marital status and residential status). Abbreviations: BMI, body mass index; SHS, subjective health scores; GDS, scores of the geriatric depression scale; SWB, subjective well-being; SSES, subjective socio-economic scores.

### 4.3 Cognitive alternations among the Older Population under the Digital Divide

Table 2 presents the differences of multi-domain cognitive function between DD group and ODD group. The results indicated that the only the SDMT score of processing speed (F=10.67, *p*<0.001) and the working memory is significant between two groups. However, the effect size of processing speed is the largest (Cohen’s d=0.92).

**Table 2.**
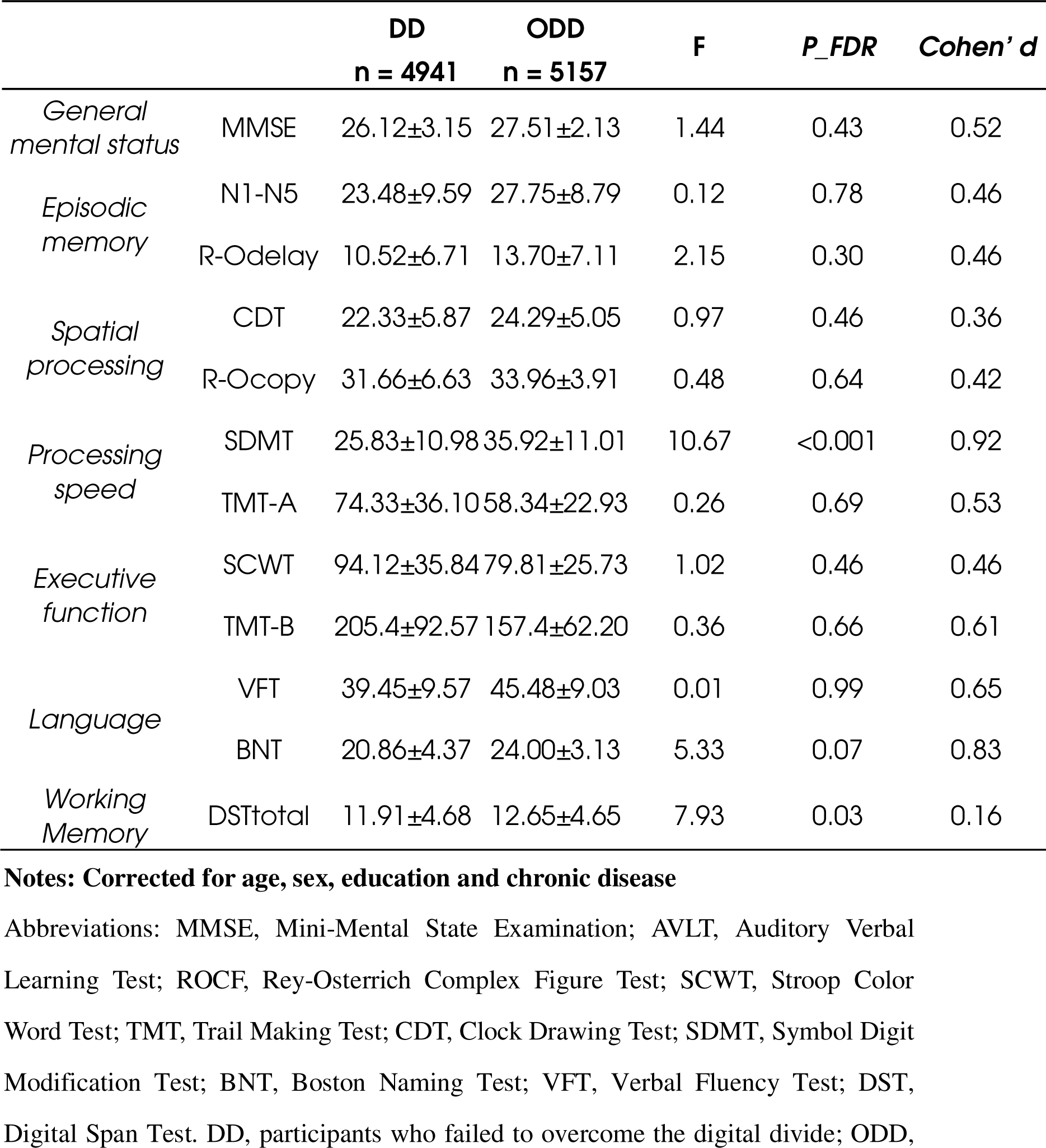
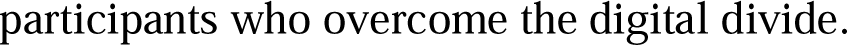
Digital divide and the cognitive performance.

### 4.4 The cross-sectional association of overcoming Digital Divide and risk of incidence of MCI

The risk of developing MCI is significantly increased in DD group compared to ODD group (See Supplemental Table 1): OR=3.06 [95% CI, 2.74-3.42]. Compared to other factors influencing MCI: age OR=1.38 [95% CI, 1.24-1.54], Diabetes OR=1.23 [95% CI, 2.74-3.42] and Hyperlipidemia OR=0.74 [95% CI, 0.66-0.83], DD has the highest odds ratio value.

### 4.5 Longitudinal evidence on the digital divide influencing cognitive aging

Significant inter-individual variations were observed for all cognitive abilities in the null model, suggesting the viability of constructing the subsequent mixed linear model (See Supplemental Table 2). However, in the unconditional growth model, only the age-related change trend of CDT did not reach statistical significance, preventing the construction of the full model (also see Supplemental Table 2).

Table 3 indicates that digital divide could significantly influenced the aging rate of multi-domain cognitive function. Compared to the ODD group and trans group, cognition of DD group displayed a faster aging rate, including MMSE (B_13_ = 3.50, *p*<0.001; B_14_ = 3.13, *p*=0.002), processing speed (TMTA: B_13_ = −1.98, *p*=0.04; B_14_ = −2.62, *p*<0.001), language (BNT: B_13_ = 3.16, *p*=0.002).

**Table 3.**
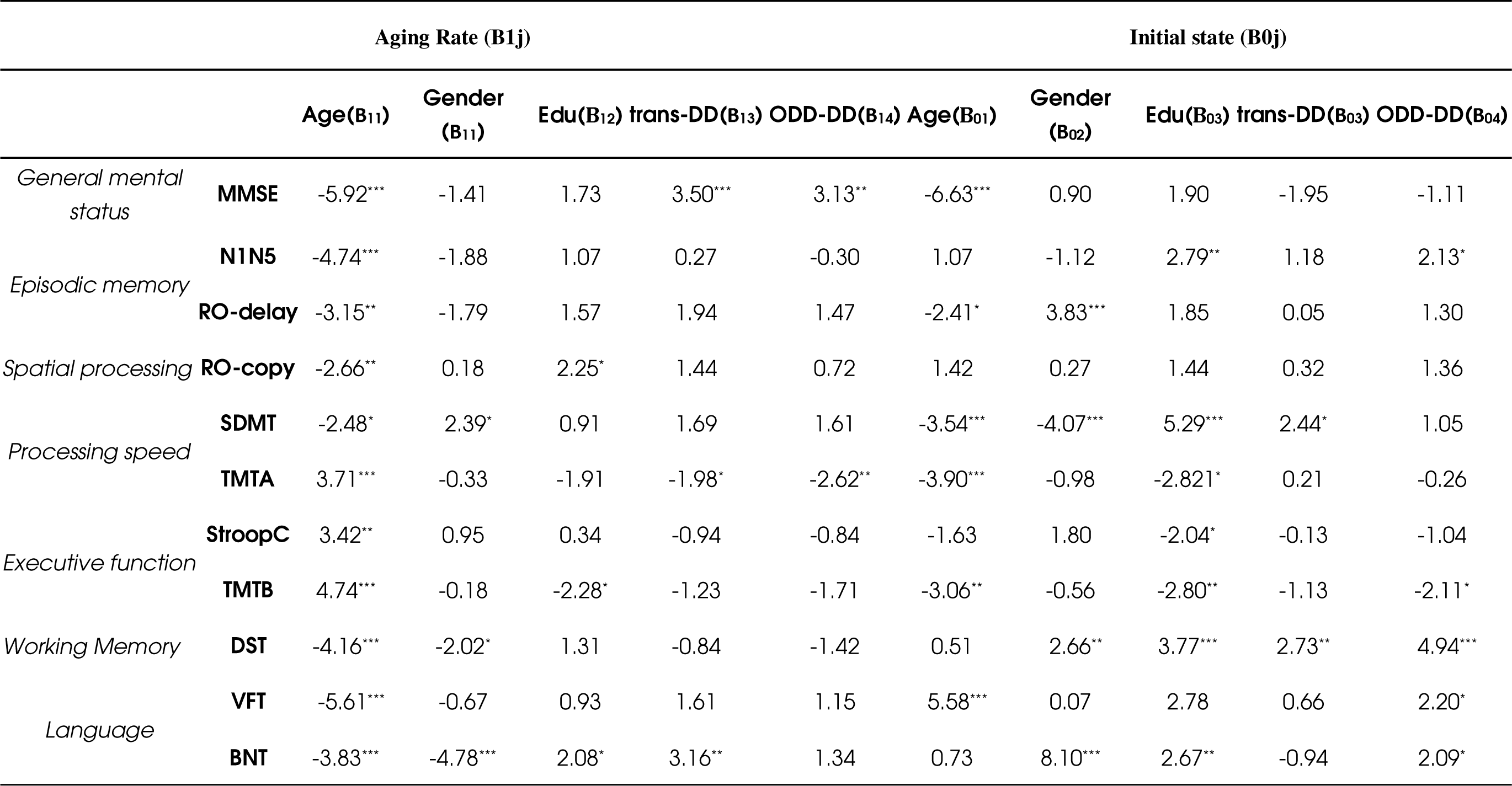
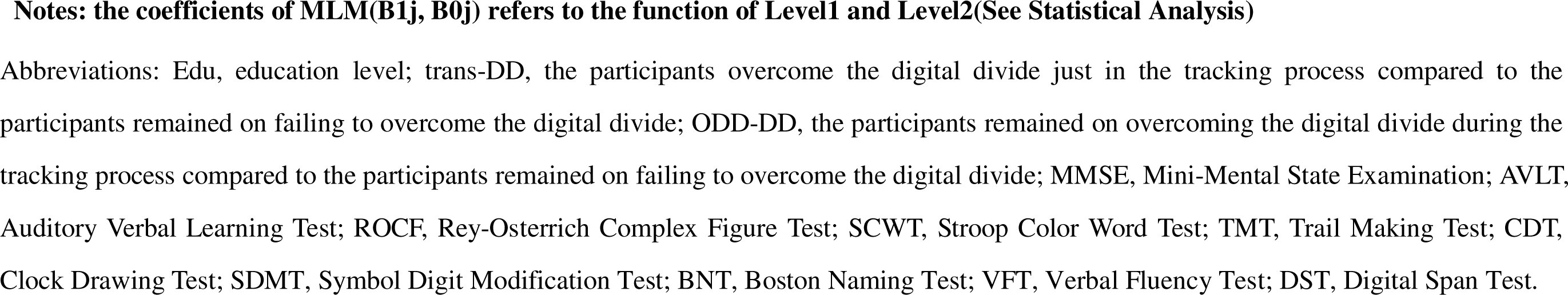
Longitudinal evidence on the digital divide influencing cognitive aging.

### 4.6 Longitudinal evidence on the digital divide influencing the development and reversion of MCI

The Table 4 indicated that compared to DD group, trans group and ODD group significantly decrease the probability of developing MCI (trans-DD: HR, 0.50; 95% CI, 0.34-0.74; ODD-DD: HR, 0.43; 95% CI, 0.29-0.62), while they have a significantly greater probability of reversion from MCI to the healthy (trans-DD: HR, 6.00; 95% CI, 3.77-9.56; ODD-DD: HR, 9.22; 95% CI, 5.63-15.11).

**Table 4.**
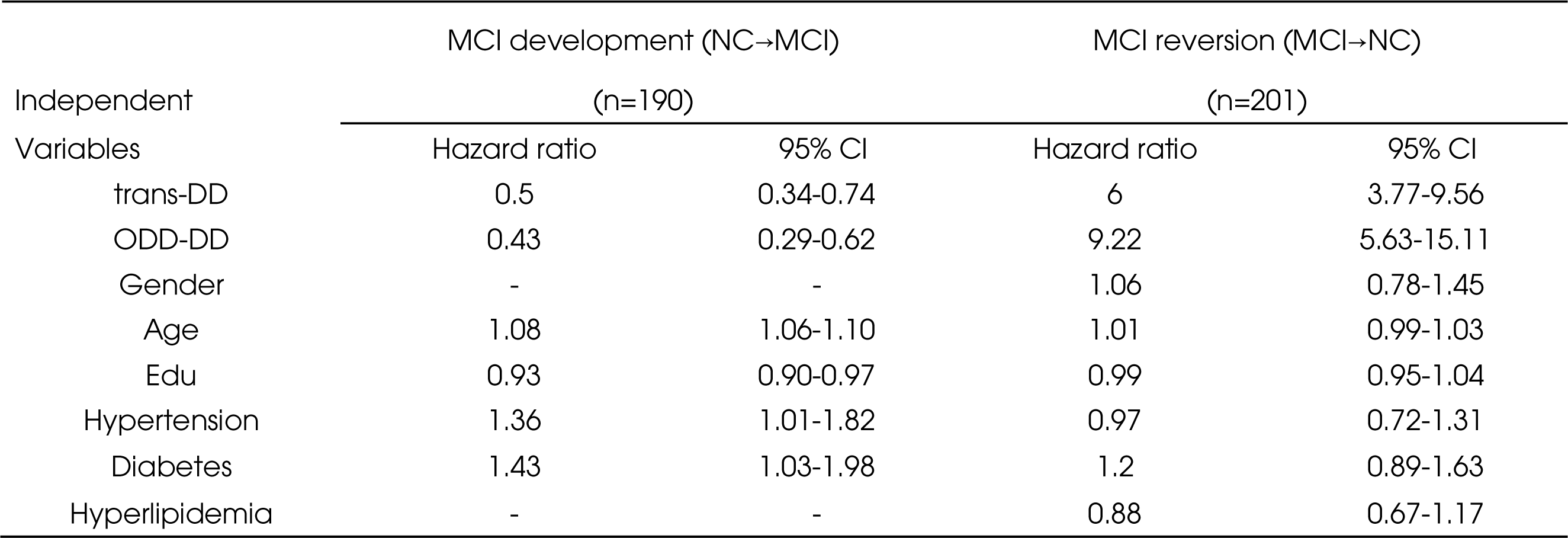
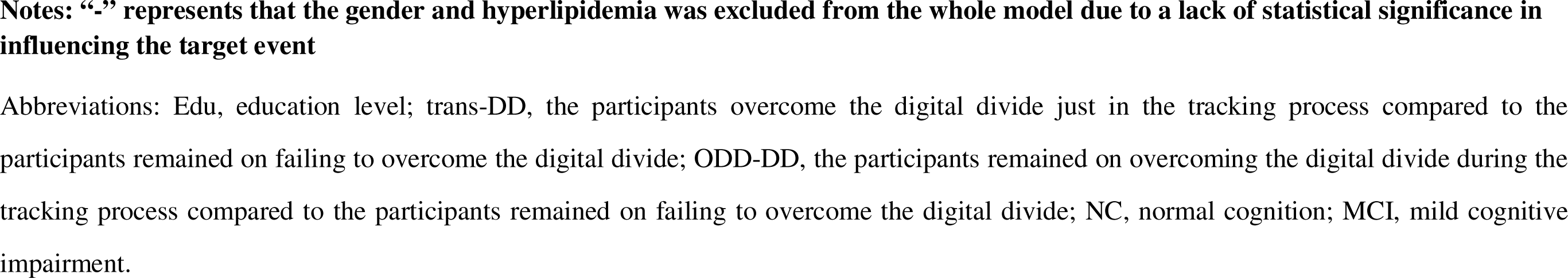
Longitudinal evidence on the digital divide influencing the development and reversion of MCI.

| Independent Variables | MCI development (NC→MCI)<br>(n=190) |  | MCI reversion (MCI→NC)<br>(n=201) |  |
| --- | --- | --- | --- | --- |
|  | Hazard ratio | 95% CI | Hazard ratio | 95% CI |
| trans-DD | 0.5 | 0.34-0.74 | 6 | 3.77-9.56 |
| ODD-DD | 0.43 | 0.29-0.62 | 9.22 | 5.63-15.11 |
| Gender | - | - | 1.06 | 0.78-1.45 |
| Age | 1.08 | 1.06-1.10 | 1.01 | 0.99-1.03 |
| Edu | 0.93 | 0.90-0.97 | 0.99 | 0.95-1.04 |
| Hypertension | 1.36 | 1.01-1.82 | 0.97 | 0.72-1.31 |
| Diabetes | 1.43 | 1.03-1.98 | 1.2 | 0.89-1.63 |
| Hyperlipidemia | - | - | 0.88 | 0.67-1.17 |

**Notes: “-” represents that the gender and hyperlipidemia was excluded from the whole model due to a lack of statistical significance in influencing the target event**
Abbreviations: Edu, education level; trans-DD, the participants overcome the digital divide just in the tracking process compared to the participants remained on failing to overcome the digital divide; ODD-DD, the participants remained on overcoming the digital divide during the tracking process compared to the participants remained on failing to overcome the digital divide; NC, normal cognition; MCI, mild cognitive impairment.

### 4.7 Sensitivity analyses

In order to verify that the cross-group differences between the DD group and the ODD group were not due to differences in age and education level, we randomly selected 50% of the participants from the DD group. We used the Propensity Score Matching (PSM) method to match these participants with the ODD group based on age, education level, and gender. We conducted the same statistical analysis using identical procedures, and the results were as follows. The horizontal results were largely replicated (See Supplemental Table 3-5).

## 5 Discussion

This population-based cohort study has been verified the causal pathway of the digital divide driven by resources inequality resulting from various influencing factors. It also revealed a significant association between the digital divide and both cross-sectional and longitudinal differences in cognition, as well as the development and reversion of MCI. The population that overcome the digital divide exhibited higher scores in processing speed compared to those who suffered from the digital divide. Also, overcoming the digital divide was associated with a slower aging rate in MMSE, processing speed and language skills, along with a reduced probability of developing MCI and an increased probability of transition from MCI into a healthy state.

The current study examines, for the first time, the impact of the digital divide on multiple cognitive domains and reversion of MCI in the aging population. Additionally, a causal model of the digital divide among the elderly has been developed. Due to its correlation with the use of social media, previous studies have often associated it with mental health issues.^22,42,43^ Nonetheless, our work aims to shift public attention to the cognitive abilities that are essential in supporting the daily functioning of aging population, in response to the potential disability that may arise during their aging process. As the results have indeed demonstrated the digital divide is significantly associated with the development and reversion of MCI. Given the numerous clinical treatment failures observed in Alzheimer’s disease (AD),^44,45^ MCI has increasingly been recognized as a crucial opportunity for disease treatment and intervention.^46,47^ The innovative findings of our study clearly indicate that crossing the digital divide (DD) plays a significant role in the conversion from MCI to NC. Despite a tracking period of only 2-3 years, the conversion rate is more than six times higher than DD group. This provides valuable insights for early intervention in the MCI stage of AD, since the utilization of ICTs is cost-effective and yields a significant impact on neuroplasticity. The underlying reason for its considerable benefits is that ICTs reshapes individuals’ interactive environment, exposing ICTs’ users to a wealth of environmental stimuli and information.^48–50^ This is especially applicable to older adults because bridging the digital divide exposes them to a new lifestyle characterized by abundant information, necessitating a learning process. Consistent with previous studies on the purpose of online cognitive training to improve cognitive function in older adults, providing further epidemiological evidence for neural plasticity and cognitive improvement during the aging process, suggesting the development of cognitive training methods integrated with digital platforms and portable devices.

In addition, our findings also indicate that the impact of the digital divide on different cognitive functions is selective. Overcoming the digital divide, both at the cross-sectional group level and longitudinal developmental level, has a significant positive effect on processing speed. The continuous influx of online information and the multimedia streams with multiple presentation modes during ICTs’ usage encourage older adults to engage in attention switching and simultaneous attention to multiple tasks, thereby enhancing the processing speed of cognitive resources.^51^ Furthermore, studies have shown that many cognitive-related variables are considered to reflect processing speed, especially in the aging process.^52^ Therefore, the influence of the digital divide on processing speed is also most evident in overall cognitive abilities (MMSE) according to our findings.

### 5.1 Limitations

First, during the third wave of longitudinal data collection in this study, the sample size of the data was lower than expected due to the COVID-19 pandemic. Second, variables used to measure the mental health were relatively few, which may not fully capture the psychological factors of the participants. Third, regarding the quantification of the digital divide, the current study did not distinguish the specific uses of ICTs. It is possible that some participants passively received phone calls and messages without actively engaging with ICTs to receive more stimuli. They may have been misclassified into the group assigned to the ODD group.

## 6 Conclusion

In this cohort study, the underlying mechanism of the digital divide among elderly individuals is influenced by their personal physiological and psychological characteristics, as well as their positional status, impacting their access to economic, cognitive, and social resources, which ultimately determines their ability to overcome the digital divide. Based on the cross-sectional association, the effect size of processing speed is the most prominent by digital divide. Meanwhile, overcoming the digital divide could delay the decline of general cognition, processing speed and part of language function. Overcoming the digital divide also decreased the development of MCI and increased the reversion of MCI. These findings can help inform strategies for dementia prevention and cognitive reserve strengthening in later life, in the context of modifiable daily routines.

## Data Availability

Raw data of the Beijing Aging Brain Rejuvenation (BABRI)
Project are available from the corresponding author on reasonable request. Restriction of raw data is to protect the privacy of participants.

## Acknowledgments and Funding Disclosure

We are grateful to all the participants and their relatives as well as our cooperating community for helping us complete this trial. This work was supported by STI2030-Major Projects (2022ZD0211600), the Natural Science Foundation of China (grant number 32171085), State Key Program of National Natural Science of China (grant number 82130118), the Fundamental Research Funds for the Central Public welfare research institution(ZZ13-YQ-073, Z0601) and Tang Scholar.

## Supplementary Materials

**Supplemental Table 1.** The cross-sectional association of overcoming Digital Divide and risk of incidence of MCI.

|  | OR | 95%CI | *P* |
| --- | --- | --- | --- |
| **Digital Divide** |  |  |  |
| ODD | 1 | (2.74 - 3.42) | <.001 |
| DD | 3.06 |  |  |
| **Age** |  |  |  |
| < 65 year | 1 | (1.24 - 1.54) | <.001 |
| > 65 year | 1.38 |  |  |
| **EDU** |  |  |  |
| College degree | 1 | (1.08 – 2.12) | 0.017 |
| Non-college | 1.51 |  |  |
| **Hyperlipidemia** |  |  |  |
| No | 1 | (0.66 – 0.83) | <.001 |
| Yes | 0.74 |  |  |
| **Diabetes** |  |  |  |
| No | 1 | (1.08 – 1.40) | 0.002 |
| Yes | 1.23 |  |  |
| **Hypertension** |  |  |  |
| No |  |  | 0.079 |
| Yes |  |  |  |

Notes: The Hypertension was excluded from the whole model due to a lack of statistical significance in influencing the target event

**Supplemental Table 2.**
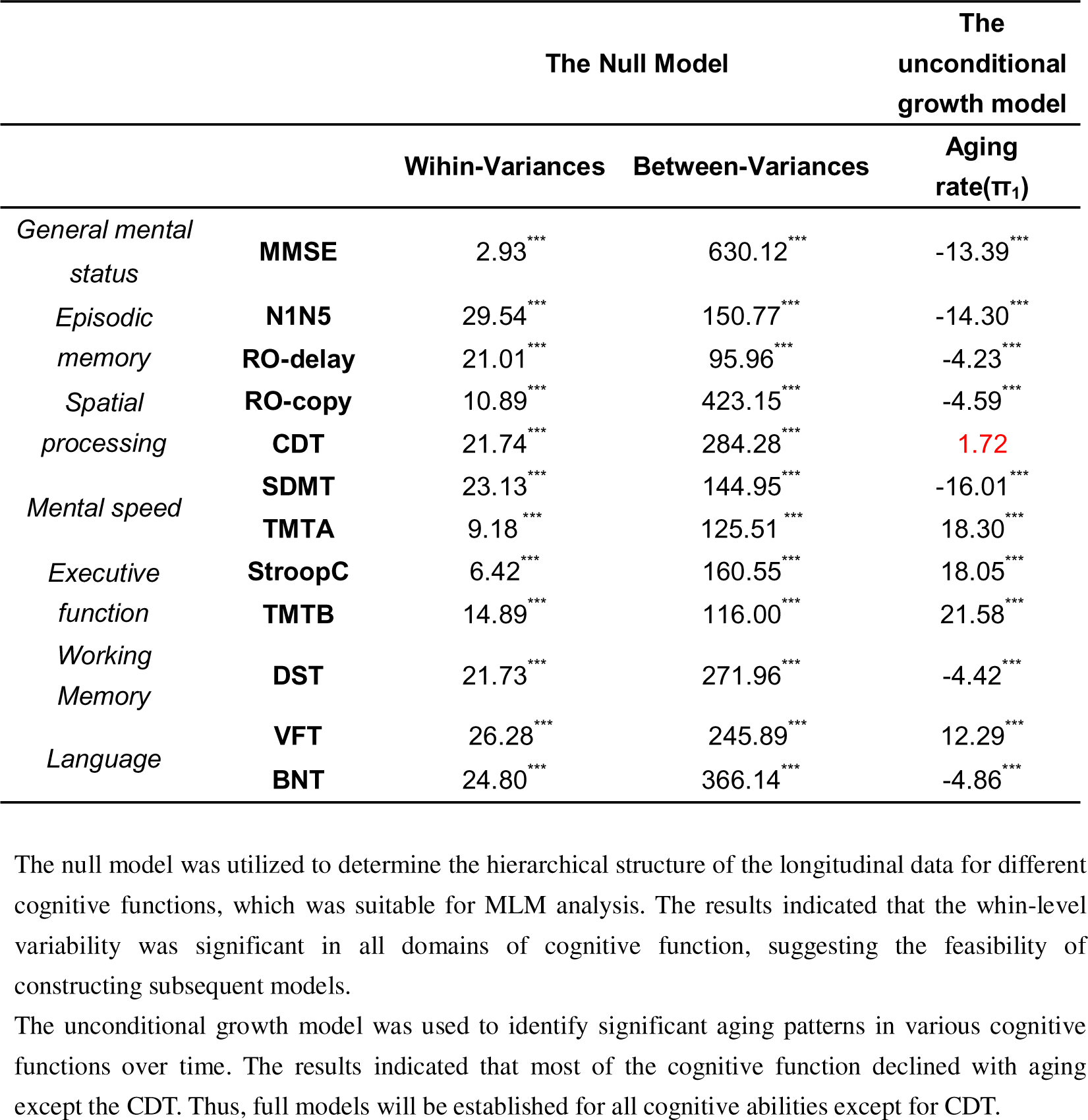
The cross-sectional association of overcoming Digital Divide and risk of incidence of MCI.

**Supplemental Table 3.**
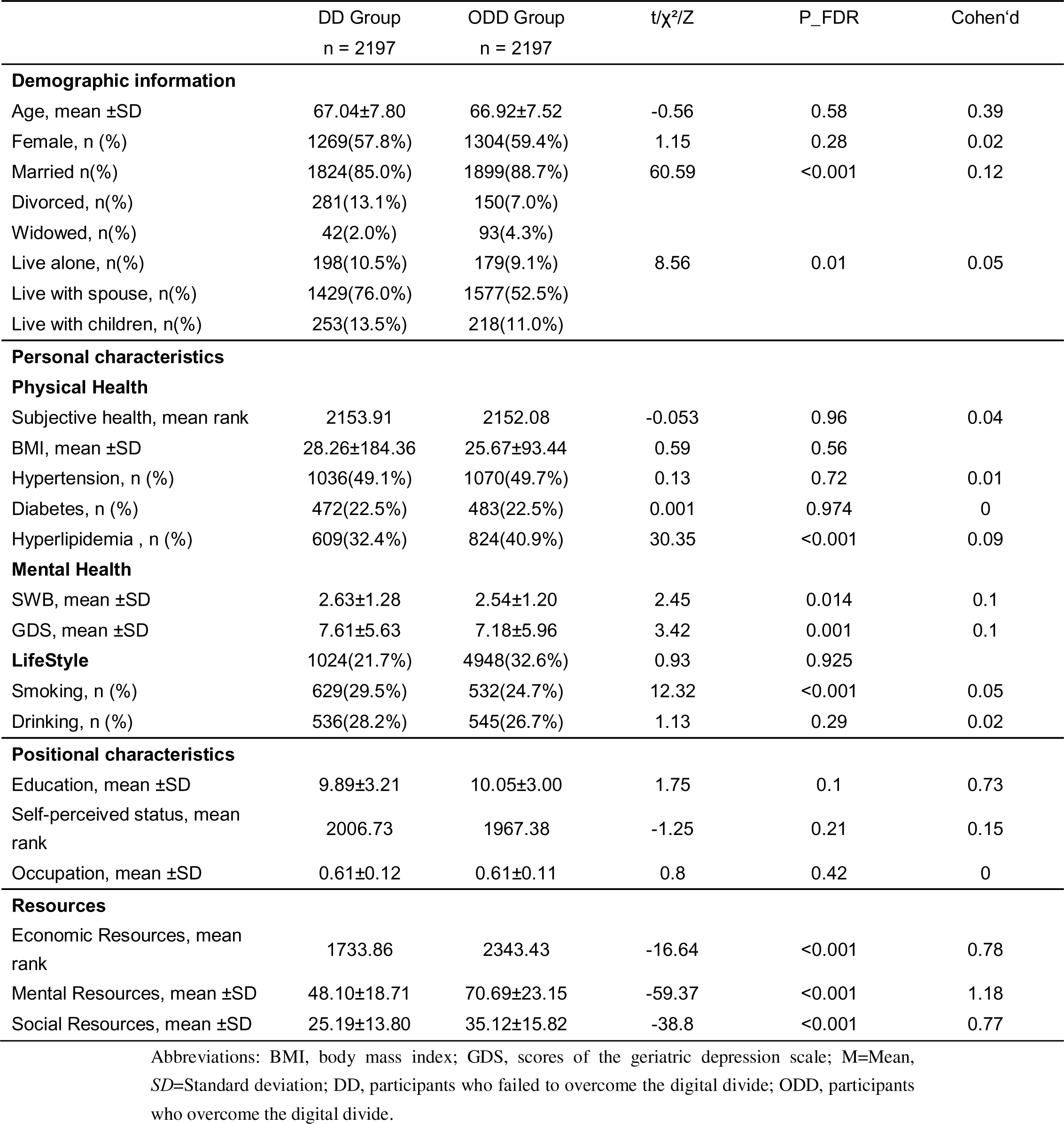
Characteristics and Digital Divide among older population (After PSM)

**Supplemental Table 4.** ANCOVA: digital divide and the cognitive performance (After PSM)

|  | **DD** | **ODD** |  |  |
| --- | --- | --- | --- | --- |
|  | **n = 2197** | **n = 2197** | **F/$\chi^2$** | **P_FDR** |
| MMSE | 26.36±3.00 | 27.23±2.33 | 2.24 | 0.13 |
| N1-N5 | 24.12±9.58 | 26.37±8.98 | 0.12 | 0.73 |
| RO-delay | 10.75±6.63 | 12.81±6.94 | 2.06 | 0.15 |
| CDT | 22.53±5.78 | 23.94±5.20 | 0.53 | 0.47 |
| RO-copy | 31.99±6.34 | 33.46±4.54 | 0.69 | 0.41 |
| SDMT | 27.02±10.72 | 32.70±10.46 | 3.97 | 0.04 |
| TMT-A | 71.70±33.91 | 62.09±24.61 | 1.55 | 0.21 |
| StroopCTime | 91.99±33.97 | 83.46±26.60 | 2.46 | 0.12 |
| TMT-B | 198.53±86.51 | 171.77±69.48 | 1.09 | 0.3 |
| VFT | 39.95±9.39 | 44.05±8.80 | 2.52 | 0.11 |
| BNT | 21.15±4.18 | 23.41±3.42 | 0.39 | 0.53 |
| DSTtotal | 11.22±2.19 | 12.23±4.47 | 1.69 | 0.19 |

**Notes: Corrected for age, sex, education and chronic disease**
Abbreviations: MMSE, Mini-Mental State Examination; AVLT, Auditory Verbal Learning Test; ROCF, Rey-Osterrich Complex Figure Test; SCWT, Stroop Color Word Test; TMT, Trail Making Test; CDT, Clock Drawing Test; SDMT, Symbol Digit Modification Test; BNT, Boston Naming Test; VFT, Verbal Fluency Test; DST, Digital Span Test. DD, participants who failed to overcome the digital divide; ODD, participants who overcome the digital divide.

**Supplemental Table 5.** The cross-sectional association of overcoming Digital Divide and risk of incidence of MCI (After PSM)

|  | OR | 95%CI | P |
| --- | --- | --- | --- |
| **Digital Divide** |  |  |  |
| ODD | 1 | (1.82 - 2.50) | <.001 |
| DD | 2.14 |  |  |
| **Age** |  |  |  |
| < 65 year | 1 | (1.23 - 1.69) | <.001 |
| > 65 year | 1.44 |  |  |
| **EDU** |  |  |  |
| College degree | 1 | (1.48 – 2.10) | <.001 |
| Non-college | 1.76 |  |  |
| **Hyperlipidemia** |  |  |  |
| No | 1 | (0.62 – 0.88) | <.001 |
| Yes | 0.74 |  |  |
| **Diabetes** |  |  |  |
| No | 1 | (1.01 – 1.48) | 0.04 |
| Yes | 1.22 |  |  |
| **Hypertension** |  |  |  |
| No | 0.99 | (0.85-1.17) | 0.94 |
| Yes |  |  |  |

